# Characteristics of Patients with Hematologic Malignancies Without Seroconversion Post-COVID19 Third Vaccine Dosing

**DOI:** 10.1101/2022.04.07.22273430

**Authors:** Sigrun Hallmeyer, Michael A. Thompson, Veronica Fitzpatrick, Yunqi Liao, Michael P. Mullane, Stephen C. Medlin, Kenneth Copeland, James L. Weese

## Abstract

Patients with hematologic malignancies (HM) are at greater risk of severe morbidity and mortality caused by COVID19 and show a lower response to a two-dose COVID19 mRNA vaccine series. The primary objective of this retrospective cohort study is to explore the characteristics of the subset of patients with HM who had little to no change in SARS-CoV-2 spike antibody titer levels after a 3rd vaccine dose (3V) (-/-). As a secondary objective, we seek to compare the cohorts of patients who did and did not seroconvert post-3V to get a better understanding of the demographics and potential drivers of serostatus. A total of 625 patients with HM had two titer results at least 21 days apart pre- and post- the 3V dose. Among the participants who were seronegative prior to 3V (268), 149 (55.6%) seroconverted after the 3V dose and 119 (44.4%) did not. HM diagnosis was significantly associated with seroconversion status (P = .0003) with patients non-Hodgkin lymphoma 6 times the odds of not seroconverting compared to multiple myeloma patients (P = .0010). Among the cohort of patients who remained seronegative post-3V, 107 (90.0%) patients showed no reaction to 3V as indicated by pre- and post- 3V index values. This study focuses on an important subset of patients with HM who are not seroconverting after the COVID mRNA 3V, providing much needed data for clinicians to target and counsel this subset of patients.

## Introduction

Patients with hematologic malignancies (HM) are at greater risk of severe morbidity and mortality caused by COVID19.^1,2^ Patients with HMs who are older, with more comorbidities, specific HM conditions, response statuses, and therapies are even more likely to experience adverse health effects caused by COVID19, including anywhere from 25% to 34% increased risk of death.^3,4^ This is believed to be due the high levels of immunosuppression making the effects of COVID19 more severe.^3^ Among HM conditions, patients with myeloid malignancies seem to be at highest risk for COVID19 associated morbidity and mortality, but this varies greatly based on treatment type.^2-6^

Moreover, patients with HMs show a lower response to the standard two-dose COVID19 mRNA vaccine series when compared to individuals with other immune suppressing conditions leaving them still more vulnerable to COVID19 associated morbidity and mortality to their non-HM counterparts.^7-9^ A recently published meta-analysis put the pooled proportion of patients achieving a serologic response after a second dose of the vaccine around 87% but was sure to acknowledge that patients with HMs are less likely to mount a response as compared with patients with solid organ cancers (63.7% vs 94.9%), likely due to the type of HM and/or current treatments.^10,11^ Another large study, the CAPTURE study, showed that in patients with HMs, neutralizing antibodies, our current indication of vaccine effectiveness, are significantly reduced, and this appears to hold true with the most recent variant of concern (VOC), Omicron.^1,4,12,^

In the summer of 2021, when the “booster dose” was authorized under the United States Food and Drug Administration (FDA) Emergency Use Authorization (EUA), cancer patients were rightly prioritized for this third shot (3V). A third dose of the mRNA vaccine (‘3V’) has shown to be particularly beneficial for HM patients given the reduced response to the standard two dose series.^13^ With the increased likelihood of seroconversion among individuals who may have been less responsive to the standard series there is added protection against COVID19 associated morbidity and mortality with 3V; however, research is emerging to show that this may not be true for all HM conditions and/or underlying treatments and responses.

This current study builds off a previous study conducted by the same team that assessed the characteristics of a large cohort of HM patients who had received a 3V dose of the mRNA COVID19 vaccine.^9^ The primary objective of this study is to explore the characteristics of the subset of patient who had little to no change in SARS-CoV-2 spike protein titer levels post 3V (-/-) and compare to the cohort of patients who did seroconvert (-/+) to get a better understanding of the drivers of serostatus.

## Methods

This retrospective cohort study analyzed patient data on SARS-CoV-2 spike IgG antibody titers pre- and post-3V across a large Midwestern healthcare system, which consists of 26-hospitals and over 500 sites of care. the ADVIA Centaur sCOVG assay was used to provide semi-quantitative (index value) results for the detection of IgG antibodies to the receptor-binding domain (RBD) of the S1 spike antigen of the SARS-CoV-2 virus. The assay received Emergency Use Authorization from the FDA and its performance characteristics were validated internally by ACL Laboratories. Assay sensitivity (positive result agreement) was 95.5% and specificity (negative result agreement) was 99.9%. An index value > 1.00 is considered positive for SARS-CoV-2 IgG antibodies.^14^

### Participants

This study included 625 patients who had been previously diagnosed with HM within the healthcare system between October 31, 2019 and January 31, 2022 and had received the full dose of a COVID19 mRNA vaccine, as defined by the CDC, two or more weeks prior to 3V.^15^ Post 3V titers were obtained 21 days after 3V. Patients were excluded from the study if vaccine status was unknown or considered incomplete, or they had received a primary series vaccination that was not an mRNA vaccine manufactured by Moderna and/or Pfizer-BioNTech. Subjects were identified via Epic EMR database by the research analytics team and de-identified to the study team. This study was determined by IRB to be non-human subjects research due to the de-identification of data (#2021-214).

### Variables

Data gathered in this study included: age, race, ethnicity, COVID19 mRNA vaccine type, COVID19 infection history prior to 3V of COVID19 vaccine, days between the second and third dose of COVID19 vaccine, HM diagnosis, up to four SARS-CoV-2 IgG antibodies results between August 28, 2021 and January 31, 2022, and days between the third dose of COVID19 vaccine and each IgG result. Age values below 90 were collected as continuous and “Age 90 or older” was recoded as 90. Sex included male and female. Race was collapsed into White, Black, Asian/Pacific Islander, and multi-racial (two or more races). Ethnicity included Hispanic/Latino and non-Hispanic/Latino. Unknown values for Race and Ethnicity were removed from the analysis. COVID19 mRNA vaccine type included Pfizer-BioNTech and Moderna. Days between vaccine doses was collected as continuous. COVID19 infection history included all EMR-documented positive SARS-CoV-2 PCR test results for COVID19 infection. IgG level values “<0.50” and “>100.00” were recoded to 0.50 and 100.00, respectively.

Diagnosis was collapsed into lymphoid leukemia, non-Hodgkin lymphoma (NHL), multiple myeloma and other plasma cell neoplasms, Hodgkin lymphoma, myeloid leukemia, other, and multiple conditions (Table 1).

**Table 1:** HM Inclusion Groups by ICD-10.

| HM Grouping | Conditions | ICD-10 |
| --- | --- | --- |
| Lymphoid leukemia | Lymphoid leukemia | C91.X* |
|  | Other lymphoid leukemia | C91.Z |
| Non-Hodgkin lymphoma | Other specific and unspecified types of non-Hodgkin lymphoma | C85.X |
|  | Non-follicular lymphoma | C83.X |
|  | Mature T/NK-cell lymphomas | C84.X |
|  | Malignant immunoproliferative diseases and certain other B-cell lymphomas | C88.X |
|  | Follicular lymphoma | C82.X |
|  | Other specified types of T/NK-cell lymphoma | C86.X |
|  | Other mature T/NK-cell lymphomas | C84.Z |
|  | Mature B-cell leukemia Burkitt-type | C91.A |
| Multiple myeloma and plasma cell neoplasms | Multiple myeloma and malignant plasma cell neoplasms | C90.X |
| Hodgkin lymphoma | Hodgkin lymphoma | C81.X |
| Myeloid leukemia | Myeloid leukemia | C92.X |
|  | Monocytic leukemia | C93.X |
|  | Leukemia of unspecified cell type | C95.X |
|  | Other leukemias of specified cell type | C94.X |
|  | Other monocytic leukemia | C93.Z |
| Graft-versus-host disease | Graft-versus-host disease | D89.81X |
| Other | Other neoplasms of uncertain behavior of lymphoid, hematopoietic and related tissue | D47.X |
|  | Other specified disorders of white blood cells | D72.8X |
|  | Other and unspecified diseases of blood and blood-forming organs | D75.8X |
|  | Eosinophilia | D72.1X |

\*X refers to 1-9

### Statistical Methods

Descriptive statistics are reported as count (percent) for categorical variables and as median (IQR) for continuous variables. Demographics and baseline variables are also reported by IgG antibody status before/after receiving 3V dose.

To assess the association between each individual characteristics and seroconversion status, patients were placed into two groups based on IgG antibody status pre-and post-the 3V dose, (-/+) and (-/-). Categorical variables were compared using a Chi-squared test or a Fisher’s exact test, as appropriate, and continuous variables using a Mann-Whitney U test. P-value less than 0.05 as an indicator of statistical significance. For categorical variables with more than two levels, p-values were calculated for both the global and the pairwise comparisons. Odds ratios are displayed as measures of association for all categorical variables. Logistic regressions were used to measure the association between HM condition and seroconversion. Three models were run to assess significant associations. Logistic regression model 1: unadjusted model fitting seroconversion on cancer diagnosis, model 2: adjusted model fitting seroconversion on cancer diagnosis, adjusting for age, model 3: adjusted model fitting seroconversion on cancer diagnosis adjusting for age, sex, and days between dose 2 and dose 3. Achieving seroconversion (-/+) was used as the reference outcome. Male, White, non-Hispanic/Latino, receiving Pfizer-BioNTech as the 3V dose, having previous COVID19 infection, and NHL were used as reference groups for the respective variables for the OR calculation.^3^ Data management and analysis were performed by the study team using R 4.1.1 (R Core Team, 2021).

### Data Sharing Statement

For original data, please contact.

## Results

A total of 625 patients with HM had two titer results at least 21 days apart pre-and post-the 3V dose. Overall, 14 patients were excluded for not having two titers results at least 21 days apart and an additional 25 patients were excluded for not having the 3V dose between the two titers. Four patients were excluded from the Race category and seven patients were excluded from the Ethnicity category for missing/unknown values. The median (range) age was 71 (64-78) years, 323 (51.7%) were male, 578 (93.1%) were White, and 605 (97.9%) were non-Hispanic/Latino (Table 2). Three hundred ninety-one (62.6%) received the Pfizer-BioNTech and 234 (37.4%) received Moderna vaccine as the 3V dose. A total of 21 (3.4%) patients had a COVID19 infection prior to receiving the 3V dose. The median (range) days between second and third doses of vaccine was 199 (177 - 224). In the overall sample, the top three conditions were 28% were diagnosed with NHL, 18.2% diagnosed with LL, 12.2% diagnosed with multiple myeloma (Table 2).

**Table 2.**
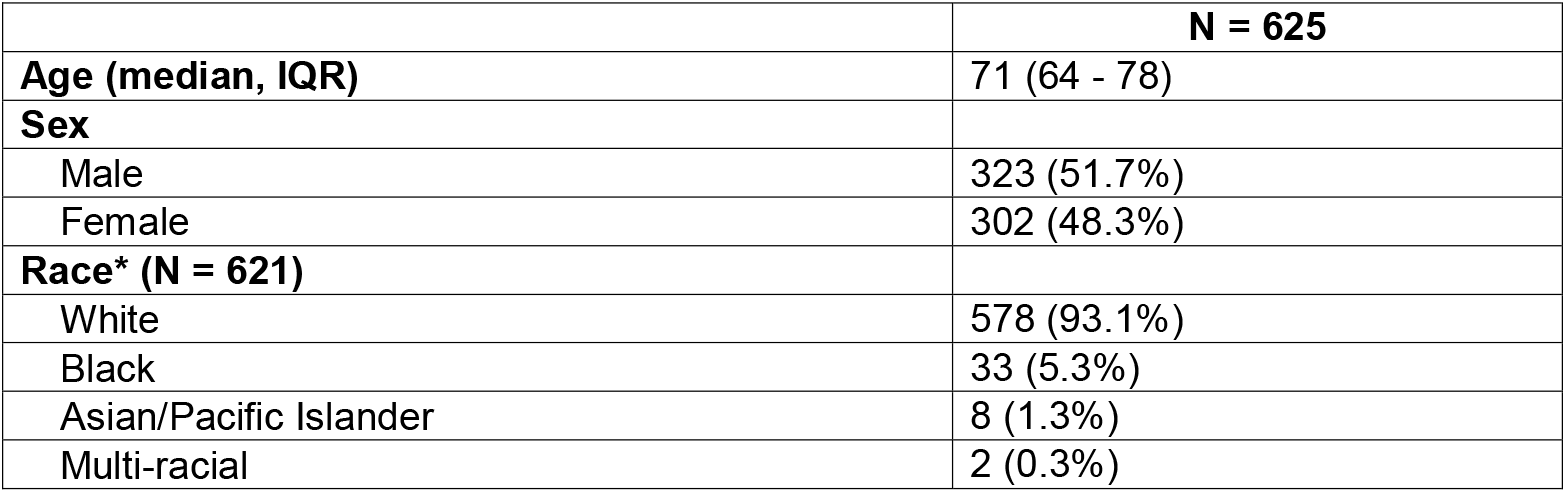
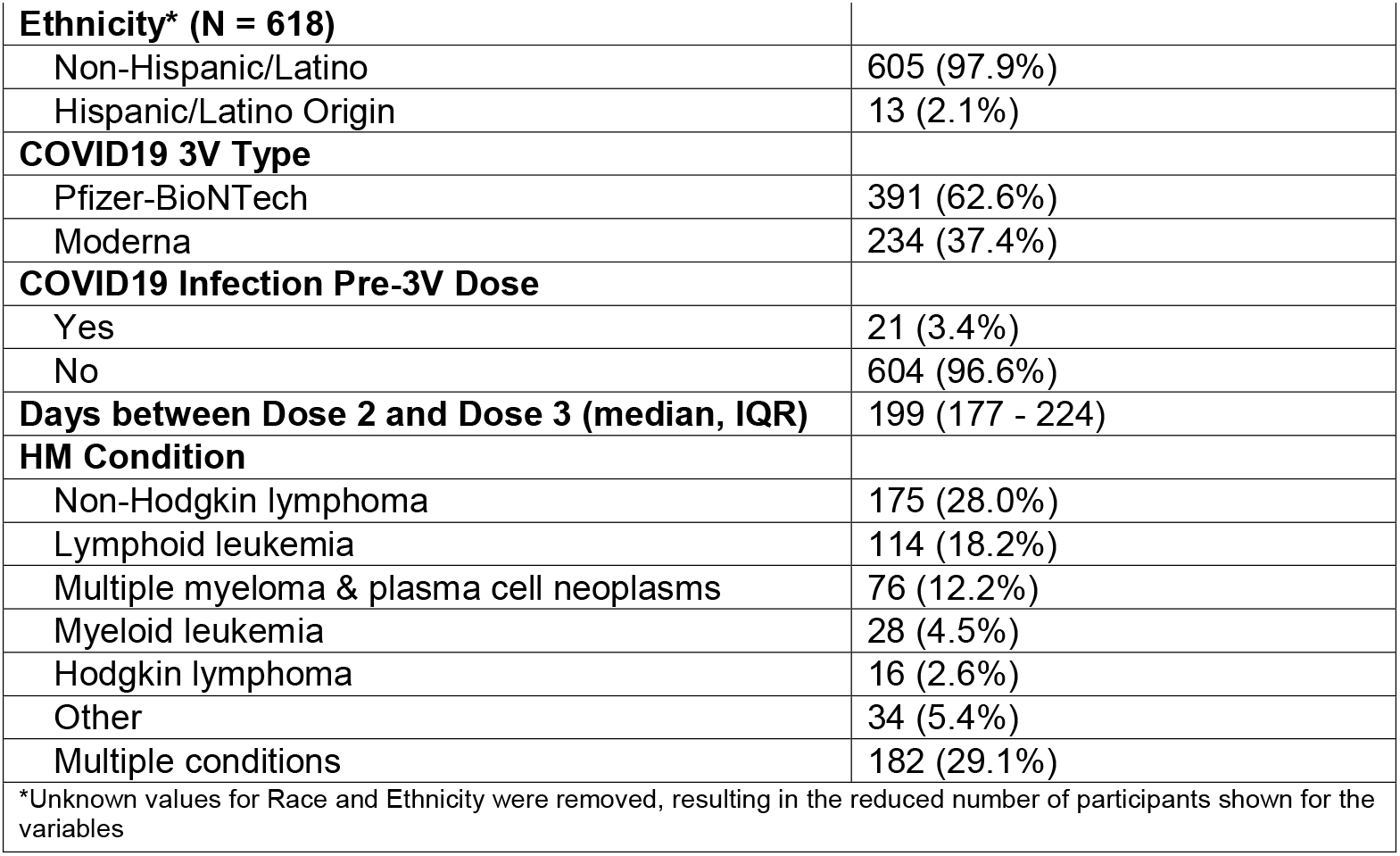
Patient Demographics.

### Comparisons by Seroconversion Status

The following analyses are only comparing the subgroup of HM patients who had a negative serostatus pre-3V, given the lower likelihood of seroconversion as compared with non-HM counterparts. Among the participants who were seronegative prior to 3V, 149 (55.6%) seroconverted after the 3V dose and 119 (44.4%) did not (Table 3). Results show the median days between dose 2 and dose 3 was significantly higher in patients who seroconverted compared to those who did not (P = .0171). Additionally, HM condition was significantly associated with seroconversion status (P = .0003). NHL patients have 6 times the odds of not seroconverting compared to multiple myeloma patients (95% CI 1.88 – 19.12, P = .0010). NHL patients also have about 14 times the odds of not seroconverting compared to patients diagnosed with the Other HM condition group (95% CI 1.72 – 112.44, P = .0021) (Figure 1).

**Table 3.**
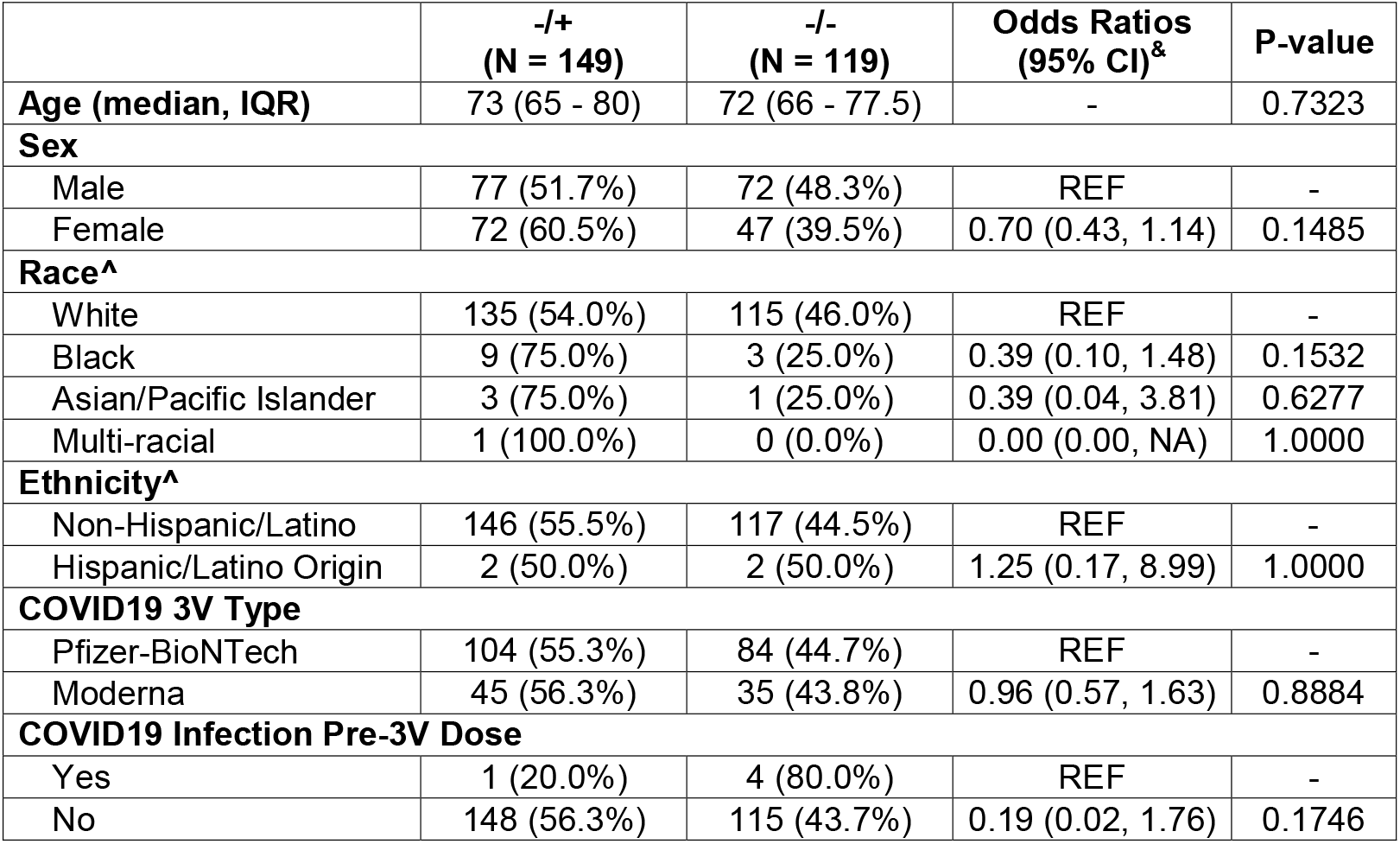
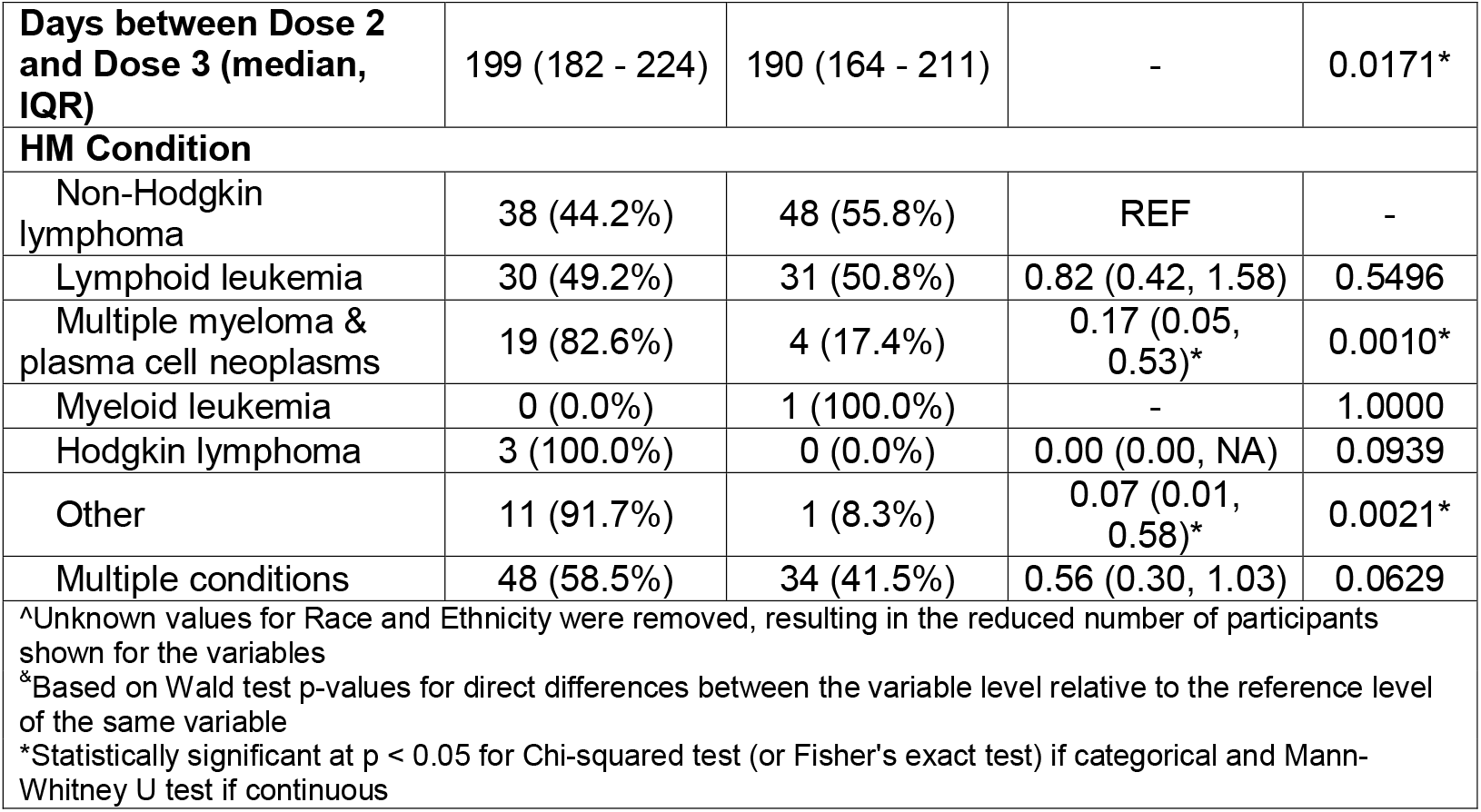
Patient Characteristics Associated with Paired IgG Results Before and After 3V Dose.

|  | **-/+**<br>**(N = 149)** | **-/-**<br>**(N = 119)** | **Odds Ratios**<br>**(95% CI)<sup>&</sup>** | **P-value** |
| --- | --- | --- | --- | --- |
| **Age (median, IQR)** | 73 (65 - 80) | 72 (66 - 77.5) | - | 0.7323 |
| **Sex** |  |  |  |  |
| Male | 77 (51.7%) | 72 (48.3%) | REF | - |
| Female | 72 (60.5%) | 47 (39.5%) | 0.70 (0.43, 1.14) | 0.1485 |
| **Race<sup>^</sup>** |  |  |  |  |
| White | 135 (54.0%) | 115 (46.0%) | REF | - |
| Black | 9 (75.0%) | 3 (25.0%) | 0.39 (0.10, 1.48) | 0.1532 |
| Asian/Pacific Islander | 3 (75.0%) | 1 (25.0%) | 0.39 (0.04, 3.81) | 0.6277 |
| Multi-racial | 1 (100.0%) | 0 (0.0%) | 0.00 (0.00, NA) | 1.0000 |
| **Ethnicity<sup>^</sup>** |  |  |  |  |
| Non-Hispanic/Latino | 146 (55.5%) | 117 (44.5%) | REF | - |
| Hispanic/Latino Origin | 2 (50.0%) | 2 (50.0%) | 1.25 (0.17, 8.99) | 1.0000 |
| **COVID19 3V Type** |  |  |  |  |
| Pfizer-BioNTech | 104 (55.3%) | 84 (44.7%) | REF | - |
| Moderna | 45 (56.3%) | 35 (43.8%) | 0.96 (0.57, 1.63) | 0.8884 |
| **COVID19 Infection Pre-3V Dose** |  |  |  |  |
| Yes | 1 (20.0%) | 4 (80.0%) | REF | - |
| No | 148 (56.3%) | 115 (43.7%) | 0.19 (0.02, 1.76) | 0.1746 |
| **Days between Dose 2 and Dose 3 (median, IQR)** | 199 (182 - 224) | 190 (164 - 211) | - | 0.0171* |
| **HM Condition** |  |  |  |  |
| Non-Hodgkin lymphoma | 38 (44.2%) | 48 (55.8%) | REF | - |
| Lymphoid leukemia | 30 (49.2%) | 31 (50.8%) | 0.82 (0.42, 1.58) | 0.5496 |
| Multiple myeloma & plasma cell neoplasms | 19 (82.6%) | 4 (17.4%) | 0.17 (0.05, 0.53)* | 0.0010* |
| Myeloid leukemia | 0 (0.0%) | 1 (100.0%) | - | 1.0000 |
| Hodgkin lymphoma | 3 (100.0%) | 0 (0.0%) | 0.00 (0.00, NA) | 0.0939 |
| Other | 11 (91.7%) | 1 (8.3%) | 0.07 (0.01, 0.58)* | 0.0021* |
| Multiple conditions | 48 (58.5%) | 34 (41.5%) | 0.56 (0.30, 1.03) | 0.0629 |
| ^Unknown values for Race and Ethnicity were removed, resulting in the reduced number of participants shown for the variables<br>*Based on Wald test p-values for direct differences between the variable level relative to the reference level of the same variable<br>*Statistically significant at $p < 0.05$ for Chi-squared test (or Fisher's exact test) if categorical and Mann-Whitney U test if continuous | | | | |

**Figure 1:**
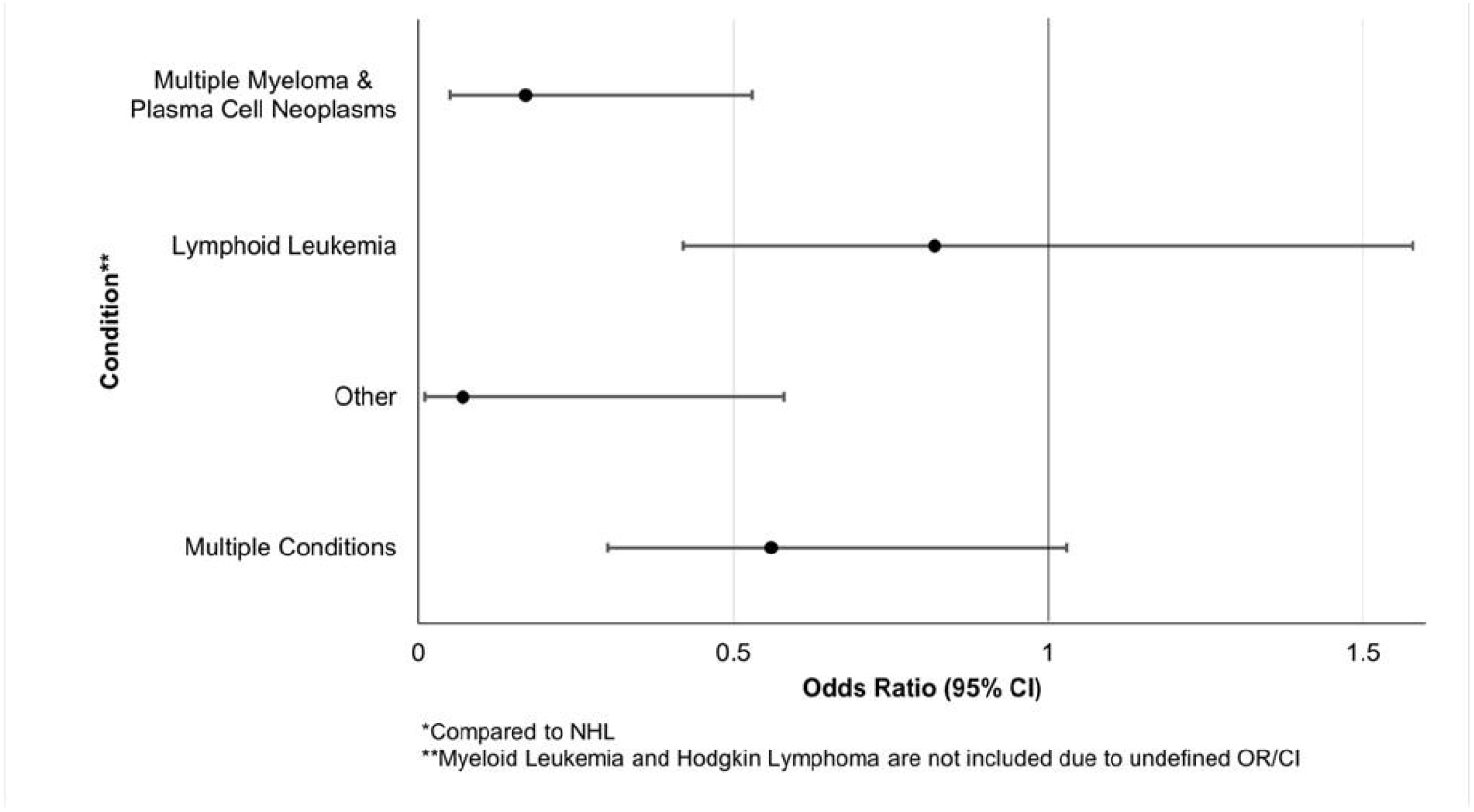
Odds Ratios by HM Condition.

Lastly, to further illustrate the relationship between HM condition and seroconversion, patients with multiple myeloma & plasma cell neoplasms or other conditions had lower odds of not seroconverting compared to NHL patients. Put conversely, NHL patients had higher odds of not seroconverting (Table 4).

**Table 4.** Models of Association Between HM Condition and Seroconversion.

|  | **Odds Ratio** | **95% CI** | **P-value** |
| --- | --- | --- | --- |
| **Model 1 (n = 268) Ref: NHL** |  |  |  |
| Lymphoid leukemia | 0.82 | (0.42, 1.58) | 0.5498 |
| Multiple myeloma & plasma cell neoplasms | 0.17 | (0.05, 0.53) | 0.0024* |
| Myeloid leukemia | - | (0, Inf) | 0.9916 |
| Hodgkin lymphoma | 0.00 | (0, Inf) | 0.9850 |
| Other | 0.07 | (0.01, 0.58) | 0.0136* |
| Multiple conditions | 0.56 | (0.30, 1.03) | 0.0638 |
| **Model 2 (n = 268) Ref: NHL** |  |  |  |
| Lymphoid leukemia | 0.81 | (0.42, 1.58) | 0.5419 |
| Multiple myeloma & plasma cell neoplasms | 0.17 | (0.05, 0.53) | 0.0025* |
| Myeloid leukemia | - | (0, Inf) | 0.9915 |
| Hodgkin lymphoma | 0.00 | (0, Inf) | 0.9848 |
| Other | 0.08 | (0.01, 0.62) | 0.0158* |
| Multiple conditions | 0.54 | (0.29, 1.01) | 0.0529 |
| Sex (Reference Group: Male) | 0.71 | (0.42, 1.18) | 0.1876 |
| Age | 0.99 | (0.97, 1.02) | 0.6267 |
| **Model 3 (n = 268) Ref: NHL** |  |  |  |
| Lymphoid leukemia | 0.84 | (0.43, 1.63) | 0.6051 |
| Multiple myeloma & plasma cell neoplasms | 0.16 | (0.05, 0.51) | 0.0022* |
| Myeloid leukemia | - | (0, Inf) | 0.9916 |
| Hodgkin lymphoma | 0.00 | (0, Inf) | 0.9851 |
| Other | 0.09 | (0.01, 0.73) | 0.0244* |
| Multiple conditions | 0.55 | (0.30, 1.02) | 0.0586 |
| Sex (Reference Group: Male) | 0.75 | (0.45, 1.27) | 0.2830 |
| Age | 1.00 | (0.98, 1.03) | 0.9724 |
| Days between Dose 2 and Dose 3 | 0.99 | (0.99, 1.00) | 0.0646 |
| *P-value significant at <0.05 |  |  |  |

### Post-3V Serostatus

Among the cohort of patients who did not seroconvert, 107 (90.0%) patients showed no reaction to 3V as indicated by pre- and post-3V index values. This subgroup had a pre-3V index value and a post-3V index value difference of <0.05 AU/ml. The additional 12 (10.0%) patients showed an average difference in pre- and post-3V of 0.086 AU/ml, indicating almost all the patients who did not seroconvert post-3V had almost no measurable antibody response to 3V using this assay. Among the cohort of patients who did seroconvert, pre-and post-3V average was 43.640, showing a much stronger response to the 3V (figure 2). Figure 2 bubbles represent weight/counts.

**Figure 2:**
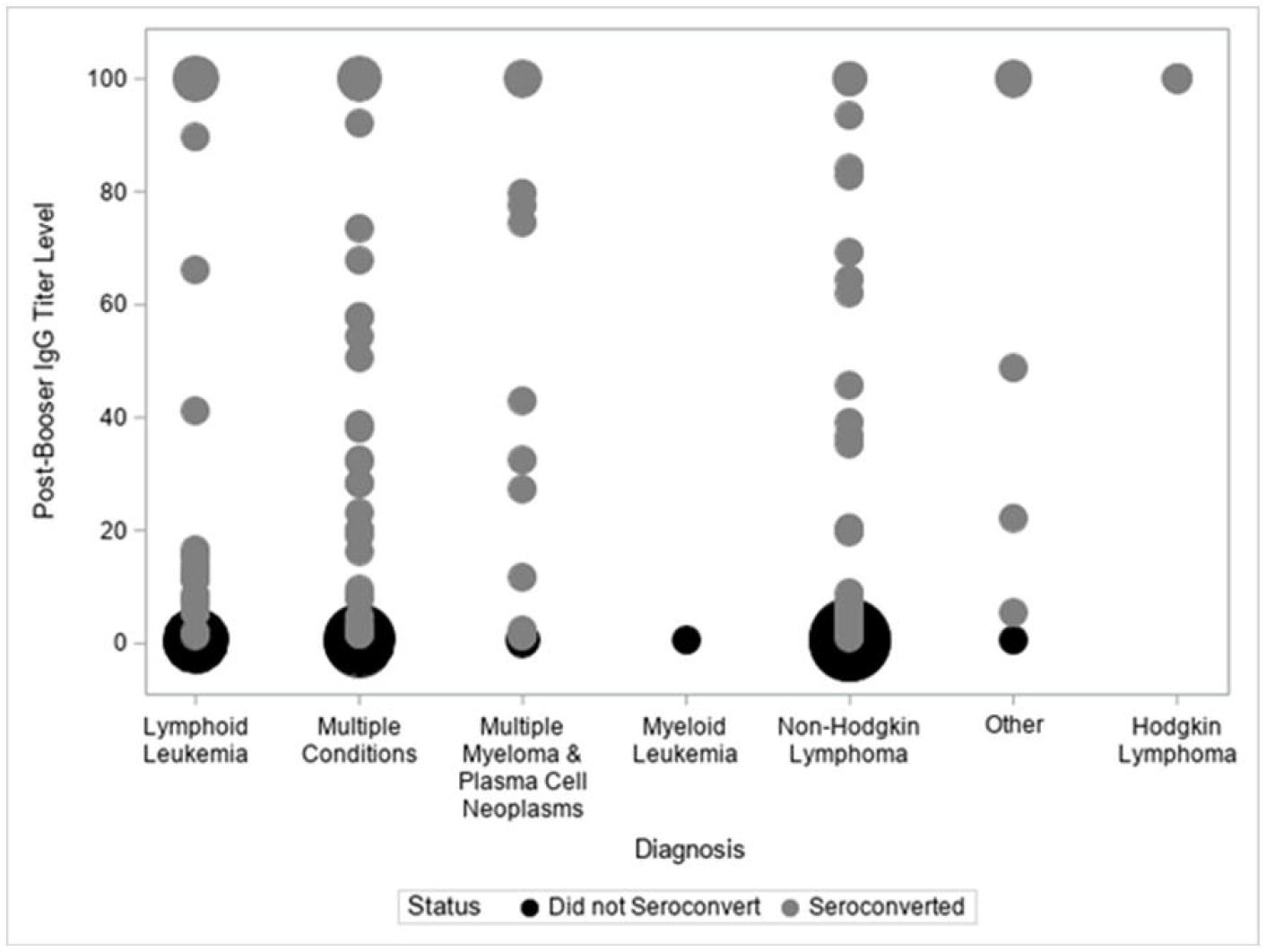
SARS-CoV-2 IgG Counts by Condition.

## Discussion

This study focuses on an important subset of HM patients who did not seroconvert after the COVID mRNA 3V. This study compares the cohort of patients who did not seroconvert (-/-) to the cohort of patients who did seroconvert (-/+) to get more insight into the differences between these two groups. Analysis shows that seroconversion is not driven by patient demographics or vaccine type. Seroconversion is however driven by HM diagnosis type and days between dose 2 and 3V – with seroconversion more likely with patients who had more days between dose 2 and 3. Although both cohorts (-/-) and (-/+) had all HM conditions represented, analysis showed that patients with NHL have 6 times the odds of not seroconverting compared to multiple myeloma patients. Additionally, NHL patients have approximately 14 times the odds of not seroconverting compared to patients diagnosed with the Other HM condition group (Table 1 for “other” conditions).

Perhaps most interesting is that the analysis showed very little to no reaction to 3V among the -/- cohort, raising interesting clinical questions about how to counsel this group of patients and whether this group may or may not benefit from a fourth dose.

### Limitations

Due to this study data being obtained from a standard of care internal program it lacked T-cell function or neutralizing antibody functional measurements. Additionally, as part of real-world data the patients were not a homogeneous population with the same treatment schedules, however this data may complement such data sets. Future studies should dive more deeply into individual HM patient characteristics as seroconversion may also be a function of disease and/or treatment.

## Conclusion

This study focuses on HM patients who are not seroconverting after the COVID mRNA 3V, suggesting a prioritized population for continued increased behavioral precautions, additional vaccination efforts, including a fourth dose of an mRNA COVID vaccine, as well as passive immunity boosting through monoclonal and or polyclonal antibodies.

## Data Availability

All data produced in the present study are available upon reasonable request to the authors

## Acknowledgments

This study received no external funding support. This study was funded by the healthcare system in which patients were used for analysis. Additionally, Christopher Blumberg and Benzon Salazar from our research analytics team supported this study by working with the research team to gather data from our EMR system. Anne Rivelli helped with data visuals, and Robert Citronberg, MD, Tulio Rodriguez, MD, Karen Gordon, Katie Cleland, and Amy Bock, helped with conceptualization and implementation of the internal clinical project in which a group of oncologists proactively asked their patients to draw titers.

## Authorship Contributions

Drs. Hallmeyer, Thompson, Medlin, Mullane, and Weese all contributed to the study conception and titer project execution. Dr. Copeland contributed to testing information. Dr. Fitzpatrick and Ms. Liao contributed to the write-up, analysis, and interpretation of data. All authors contributed to manuscript writing and review. Manuscript has been approved by all authors.

## References

1. Passamonti, F., Cattaneo, C., Arcaini, L, Bruna, R, Cavo, M, et al. Clinical characteristics and risk factors associated with COVID-19 severity in patients with haematological malignancies in Italy: a retrospective, multicentre, cohort study. The Lancet Haematology. 2020; 7 (10) E737–E745

2. Cattaneo, C., Daffini, R., Pagani, C., Salvetti, M, Mancini, V. et al. Clinical characteristics and risk factors for mortality in hematologic patients affected by COVID-19. Cancer. 2020; 126 (23) 5069–5076 https://acsjournals.onlinelibrary.wiley.com/doi/10.1002/cncr.33160

3. Garcia-Suarez, J., de la Cruz, J., Cedillo, A., Llama, P., Duarte, et al. Impact of hematologic malignancy and type of cancer therapy on COVID-19 severity and mortality: lessons from a large population-based registry study. Journal of Hematology & Oncology. 2020; 13 (133). Published online: https://jhoonline.biomedcentral.com/articles/10.1186/s13045-020-00970-7

4. Thakkar, A., Pradhan, K., Jindal, S., Cui, Z., Rockwell, B. et al. Patterns of seroconversion for SARS-CoV-2 IgG in patients with malignant disease and association with anticancer therapy. Nature Cancer. 2021; 2, 392–399 https://www.nature.com/articles/s43018-021-00191-y

5. Li, W., Wang, D., Guo, J., Yuan, G., Yang, Z., et al. COVID-19 in persons with chronic myeloid leukaemia. Leukemia. 2020; 34, 1799–1804 https://www.nature.com/articles/s41375-020-0853-6

6. Fox, T., Troy-Barnes, E., Kirkwood, A., Chan, W., Day, J. et al. Clinical outcomes and risk factors for severe COVID-19 in patients with haematological disorders receiving chemo-or immunotherapy. British Journal of Haematology. 2020; 191 (2), 194-206 https://onlinelibrary.wiley.com/doi/10.1111/bjh.17027

7. Thakkar, A., Gonzalez-Lugo, J., Goradia, N., Gali., R., Shapiro, L. Seroconversion rates following COVID-19 vaccination among patients with cancer. Cancer Cell. 2021; 39 (8), P1081-1090 https://www.cell.com/cancer-cell/fulltext/S1535-6108(21)00285-3

8. Marra, A., Generali, D., Zagami, P., Cervoni, V., Gandini, S. et al. Seroconversion in patients with cancer and oncology health care workers infected by SARS-CoV-2. Annals of Oncology. 2020; available online: https://www.annalsofoncology.org/article/S0923-7534(20)42965-5/fulltext

9. Thompson, M., Hallmeyer, S., Fitzpatrick, V., Liao, Y., Mullane, M., et al. Real World Third COVID19 Vaccine Dosing and Antibody Response in Patients with Hematologic Malignancies. Journal of Patient-Centered Research and Reviews. 2022; in review.

10. Sakuraba, A., Luna, A. and Micic, D. Serologic response following SARS-COV2 vaccination in patients with cancer: a systematic review and meta-analysis. Journal of Hematology & Oncology. 2022; 15 (15). Available: https://jhoonline.biomedcentral.com/articles/10.1186/s13045-022-01233-3

11. Gavriatopoulou, M., Terpos, E., Malandrais, P., Ntanasis-Stathopoulos, I., Briasoulis, A., et al. Myeloma patients with COVID-19 have superior antibody responses compared to patients fully vaccinated with the BNT162b2 vaccine. British Journal of Haematology. 2021; 196 (2), 356-359 https://www.ncbi.nlm.nih.gov/pmc/articles/PMC8653218/

12. Favresse, J., Gillot., C., Di Chiaro, L., Eucher, C., Elsen, M. Neutralizing Antibodies in COVID-19 Patients and Vaccine Recipients after Two Doses of BNT162b2. Viruses. 2021:13(7), 1364 https://www.ncbi.nlm.nih.gov/pmc/articles/PMC8309994/

13. Aikawa, N., De Vinci Kanda Kupa, L., Medeiros, AC., Goncalves Schahin Saad, C., Figueireso Neves Yuki, E., et al. Increment of immunogenicity after third dose of a homologous inactivated SARS-CoV-2 vaccine in a large population of patients with autoimmune rheumatic diseases. Annals of Rheumatic Disease. 2021. Available: https://pubmed.ncbi.nlm.nih.gov/34594036/

14. Siemens Healthcare Diagnostics Inc. Fact Sheet for Healthcare Providers. ADVIA Centaur SARS-CoV-2 IgG (sCOVG). August 10, 2021.

15. Centers for Disease Control and Prevention. Stay Up To Date with Your COVID-19 Vaccines. Updated March 10, 2022. Accessed February 25, 2022: https://www.cdc.gov/coronavirus/2019-ncov/vaccines/stay-up-to-date.html

